# Cardiac autonomic function during exercise and incident Parkinson’s disease

**DOI:** 10.1101/2024.11.08.24316979

**Authors:** Stefan van Duijvenboden, Julia Ramírez, Job Scheurink, Sirwan K.L. Darweesh, Michele Orini, Andrew Tinker, Patricia B. Munroe, Jos Thannhauser, Luc Evers, Joanna IntHout, Pier D. Lambiase, Bastiaan R. Bloem, Aiden Doherty, Marc A. Brouwer

## Abstract

**Objective:** To determine whether established parameters of cardiac autonomic function are associated with incident Parkinson’s disease, independent of clinical characteristics, and established autonomic prodromal features.

**Methods Population-based cohort:** study of UK Biobank participants who performed a standardized bicycle exercise test (2009-2013), followed until November 2022, and analyzed in January 2024. Heart rate increase from rest to exercise, and the decrease in heart rate from peak exercise to recovery were extracted and associated with incident Parkinson’s disease. Associations were adjusted using multivariable models consisting of clinical characteristics only and combined with prodromal autonomic features.

**Results:** 69,288 eligible participants (male 48%, mean age 56.8 [SD 8.2]) were followed for 12.5 years (median; IQR 0.3): 319 (0.5%) developed Parkinson’s disease. Median lag time to diagnosis was 9.3 years (IQR 4.4). Both heart rate increase (37.5 [SD 11.5] vs 40.8 [SD 12.4] beats/min, p < 0.001) and recovery (23.4 [SD 8.8] vs. 27.8 [SD 10.3] beats/min, p < 0.001) were significantly lower in incident cases compared to controls. After adjusting for prodromal clinical and autonomic features, heart rate recovery was independently associated with incident Parkinson’s disease, while heart rate increase was not. Specifically, a blunted heart rate lowering during recovery was associated with a 30% higher risk of incident Parkinson’s disease (HR: 1.3; 95% CI 1.1-1.4; p < 0.001 per 10 beats less recovery)

**Interpretation:** These findings suggest that cardiac autonomic dysfunction precedes clinically manifest Parkinson’s disease, and that heart rate recovery might serve as a quantitative prodromal marker.

## Introduction

Parkinson’s disease (PD) is the world’s fastest growing neurological disorder in terms of prevalence^1^. One main clinical challenge lies in earlier detection, as significant neurological degeneration has already occurred by the time PD is identified with the prevailing diagnostic approaches^2^. At present, prediction models for the early diagnosis of PD consist of predominantly non-modifiable, binary clinical characteristics (e.g. male gender, or presence of hyposmia), resulting in rather ‘static’ models^3,4^. In contrast, a more dynamic variable that could reflect quantitative changes over time, with repeated measurements during follow-up, would offer additional insights. Such a biomarker could improve early diagnosis and capture the gradual progression towards manifest PD^2,5^. In this context, electrocardiographic (ECG) parameters that focus on cardiac autonomic measures have been proposed as a potentially important tool^6,7^.

In patients with manifest PD, altered cardiac autonomic function is common^8,9^ including impaired heart rate regulation at rest and during exercise^10^. Accumulating evidence suggests that dysregulation of autonomic processes in general, and cardiac autonomic dysfunction in particular, may precede the motor symptoms of PD^6–8^. Importantly, pathological studies in PD have demonstrated a characteristic sequence of neurodegenerative events, with early involvement of the dorsal motor nucleus of the vagal nerve^11^, which regulates (cardiac) parasympathetic tone. Cardiac imaging studies have also demonstrated changes in the sympathetic nervous system in very early stages of PD^12^. This background has motivated different initiatives in population-based cohorts to search for potential prodromal autonomic markers, including heart rate regulation^13^.

Owing to differences in population characteristics, variations in choice of autonomic markers, and non-uniformity in protocols used to assess these markers, the available evidence is difficult to interpret^14–17^. Two large cohort studies focused on heart rate variability (HRV), a marker of cardiac parasympathetic tone, as a potential prodromal marker for incident PD and reported conflicting findings^14,15^. Based on differences in maximum heart rates achieved, a small controlled exercise study suggested impaired sympathetic activity as prodromal sign for PD^16^, but this could not be confirmed in a larger cohort^17^. Here, we focus on other autonomic markers, namely the increase in heart rate during exercise and its reduction during recovery, the advantage being that these markers are easily extractable, straightforward to assess, and clinically established in different settings^18^.

We therefore studied these markers in a long-term follow-up study of 69,288 UK Biobank participants who performed an exercise test, which represents the largest investigation to date on cardiac autonomic parameters and incident PD. Our specific aim was to assess whether exercise-related parameters of cardiac autonomic function during a baseline bicycle test would be associated with incident PD during long-term follow-up.

## Methods

### Participants

The UK Biobank study comprises a total of 502,364 participants, with even numbers of men and women aged 40-69 years at recruitment from 21 assessment centers across England, Wales, and Scotland, with extensive baseline and follow-up clinical, biochemical, genetic and outcome measures. The study received approval from the North West Multi-Centre Research Ethics Committee, and all participants provided informed consent at the time of enrolment (2006 – 2010). From the full cohort, 96,524 participants consented to participate in an exercise stress test with heart-rate monitoring (2009-2013). According to protocol, 13,962 participants were considered ineligible to perform the activity (https://biobank.ndph.ox.ac.uk/ukb/ukb/docs/Cardio.pdf). Another 7221 participants who did not complete the test were excluded as well. Furthermore, exclusions were based on a history of cardiovascular disease (N=4441), a diagnosis of cancer within 1 year before or after the exercise stress test (N=1526) and a history of a neurodegenerative (N=260) disorder. In particular, we focused on the exclusion of patients with PD at the time of the exercise test (N=97) using previously described diagnostic criteria (https://biobank.ctsu.ox.ac.uk/crystal/ukb/docs/alg_outcome_pdp.pdf). Thus, self-reported diagnoses were also considered, to ensure that all potential cases with PD were excluded at baseline.

In total, 69,288 of the 96,524 (71.8%) participants who provided consent to participate in the exercise test were included in the present analysis (Figure 1).

**Figure 1.**
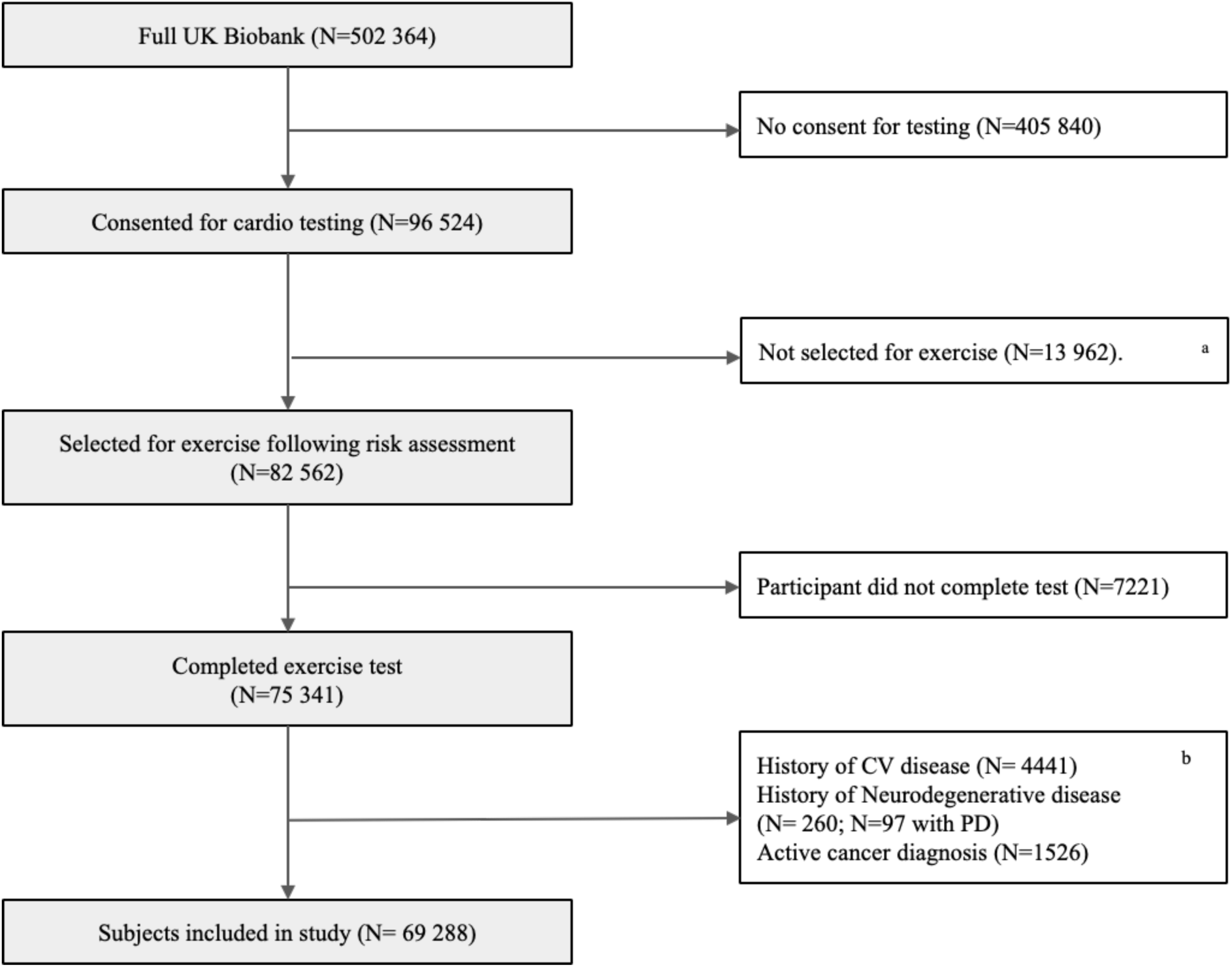
Study exclusion diagram The full list of UK Biobank variables used for disease exclusions is provided in Supplemental Table 1. ^a^ Participants ineligible to exercise had one of more safety risk factors as defined in paragraph 9.1 of the protocol manual provided by UK Biobank (https://biobank.ndph.ox.ac.uk/showcase/ukb/docs/Cardio.pdf). ^b^ Excluded subjects who met one or more of the listed criteria; CV, cardiovascular; PD, Parkinson’s disease.

### Exercise protocol

The test uses cycle ergometry on a stationary bike (eBike, Firmware v1.7) in conjunction with a single-lead ECG device (CAM-USB 6.5, Cardiosoft v6.51). According to protocol, the predicted absolute maximum workload was calculated, and participants were instructed to perform the test with a predefined target power at 35% or 50% of the maximum predicted workload (https://biobank.ndph.ox.ac.uk/ukb/ukb/docs/Cardio.pdf). In the following order, the exercise bicycle protocol consisted of: a resting phase (15 s pre-test); graded activity with increasing workload (6 min); and a recovery phase with hands remaining on the handlebars whilst remaining still and silent (1 min, no cool-down period). Heart rate measurements were available throughout the protocol.

### Cardiac Exercise Parameters of Autonomic Function

From the measurements recorded at three different phases of the exercise test we calculated 1) ‘heart rate exercise’^19^ and 2) ‘heart rate recovery’^18–20^

1. The heart rate increase during exercise (HRI-exc): heart rate at peak exercise minus resting heart rate;
2. The heart decrease during recovery (HRD-rec): heart rate at peak exercise minus recovery heart rate;

### Outcome measure & Follow-up

Diagnoses were captured using the “Spell and Episode” category from the UK National Health Service (NHS) coded according to the 9^th^ and 10^th^ revisions of the international Classification of Diseases (*ICD*-*9* and *ICD-10)*, made during hospital stay, including stays from before the study inclusion. For diagnostic validity, we refer to https://biobank.ndph.ox.ac.uk/ukb/ukb/docs/alg_outcome_pdp.pdf. Detailed information about the linkage procedure is available online (https://biobank.ndph.ox.ac.uk/ukb/ukb/docs/HospitalEpisodeStatistics.pdf). The last follow-up check was performed at 30 November 2022. Incident Parkinson’s disease was defined as ICD-10 code G20 (https://biobank.ndph.ox.ac.uk/ukb/ukb/docs/alg_outcome_pdp.pdf).

### Statistical analysis

Descriptive statistics are presented as mean (SD), median (IQR), or frequencies. Continuous data were compared using Student’s t-tests or Mann-Whitney U tests, whichever appropriate. Categorical variables were compared using either Chi-squared tests or Fisher’s exact tests. The associations between exercise parameters and incident PD were investigated using three multivariable-adjusted Cox proportional hazards regression models: 1) a minimally adjusted analysis using only age and sex as covariates to test whether exercise parameters were associated with incident PD; 2) a clinically adjusted model to address whether this association was confounded by demographical and clinical characteristics, other than established autonomic prodromal markers (model 1 + ethnic background, smoking, BMI, Townsend deprivation index, and type-2 diabetes); and 3) a fully adjusted model to evaluate whether the association had incremental value beyond other autonomic prodromal markers (constipation, depression, bladder dysfunction, and self-reported sleep duration and sleeplessness). Definitions of covariates are provided in Supplemental Table 1. Schoenfeld residuals were used to test the proportional hazards assumption, and no violation was observed. Time-to-event curves were constructed with Kaplan-Meier methods, with log-rank testing for statistical comparisons. Missing variables were imputed using the multiple imputation by chained equations approach, with five imputed datasets and ten iterations, including comparisons of distribution plots of recorded and imputed variables^21^. A P < 0·05 was considered statistically significant.

Statistical analyses were performed in R v4.2.0 using the *survival* (v3.5.8) and *mice* (v3.15.0) libraries^22^.

### Sensitivity analyses

We pre-specified sensitivity analyses to assess whether the associations found in the main analysis were affected by: (1) heart rate modulating agents, i.e. beta and calcium blockers; and (2) psychoactive drugs (excluding for example participants exposed to drugs affecting the dopaminergic system). Also, we performed analyses with and without inclusion of participants in whom data were imputed. Post-hoc sensitivity analyses were performed to further explore the associations observed in the pre-specified analyses.

## Results

In total, there were 96,524 participants who consented to partake in the exercise test, of whom 27,236 were excluded (Figure 1, Supplemental Table 2). Excluded subjects showed clinically relevant differences in baseline demographics (higher age and BMI) and in established prodromal factors of incident PD (higher prevalence of risk factors) compared to the remaining 69,288 individuals (Supplemental Table 3).

Incident PD was diagnosed in 319 individuals (0.5%) (Table 1). At the time of the exercise test, participants who developed PD during follow-up were older than those who remained disease free and were more often male.

**Table 1.** Baseline characteristics of participants with and without diagnosis of incident PD during long-term follow-up.

|  | **PD**<br>(n=319; 0.5%) | **No PD**<br>(n=68 969; 99.5%) | **N** | **p** |
| --- | --- | --- | --- | --- |
| Sex, male, % | 69.3% | 47.5% | 69 288 | <0.001 |
| Age, years , mean (SD) | 63.2 (5.2) | 56.8 (8.2) | 69 288 | <0.001 |
| Ethnicity, white, % | 95.3% | 93.1% | 68 905 | 0.13 |
| BMI, mean (SD) | 27.6 (4.0) | 27.0 (4.4) | 69 287 | 0.001 |
| Townsend deprivation index<sup>a</sup>, mean (SD) | -1.6 (2.8) | -1.4 (2.9) | 69 215 | 0.079 |
| Type 2 Diabetes Mellitus, yes, % | 13.2% | 4.2% | 69 288 | <0.001 |
| Current Smoking, yes, % | 3.2% | 8.2% | 68 977 | 0.001 |
| Constipation, yes, % | 6.0% | 3.0% | 69 288 | 0.002 |
| Depression, yes, % | 8.8% | 5.5% | 69 288 | 0.010 |
| Bladder dysfunction, % | 1.9% | 1.2% | 69 288 | 0.29 |
| Erectile dysfunction, % | 2.3% | 0.4% | 32 953 | 0.003 |
| Sleep duration, hours, mean (SD) | 7.3 (1.3) | 7.1 (1.1) | 69 182 | 0.024 |
| Sleeplessness, % | 23.1% | 25.4% | 69 035 | 0.36 |

*BMI = Body Mass Index; PD = Parkinson's disease, <sup>a</sup> Townsend deprivation index is a measure of material deprivation.*

Rates of many of the prodromal risk factors (such as constipation) were higher, while the rate of smoking (protective factor) was lower.

Median follow-up duration on survival status was 12·5 years (IQR: 0·3) with more than 10 years of follow-up available in 59,460 participants (85.6%). During this follow-up period, 3106 (4.5%) participants died.

### Heart rate response to recovery is blunted in prodromal PD

During the pre-test stage, resting heart rate was not significantly different between participants who developed PD and those who did not (Table 2). However, the increase in heart rate during exercise (HRI-exc) and decrease during recovery (HRD-rec) were both lower in the group that developed PD (Table 2).

**Table 2.** Heart-rate markers during exercise testing comparing participants with and without incident PD during long-term follow-up.

|  | **PD**<br>(n=319; 0.5%) | **No PD**<br>(n=68 969; 99.5%) | **N** | **p** |
| --- | --- | --- | --- | --- |
| **Resting Heart Rate, mean (SD)** | 70.6 (11.5) | 70.2 (11.1) | 68 103 | 0.86 |
| *HRI-exercise, mean (SD)* | 37.5 (11.5) | 40.8 (12.4) | 67 553 | <0.001 |
| **Peak Heart Rate, mean (SD)** | 107.9 (14.1) | 111.0 (13.9) | 67 780 | <0.001 |
| *HRD-recovery, mean (SD)* | 23.4 (8.8) | 27.8 (10.3) | 67 109 | <0.001 |
| **Heart Rate at Recovery, mean (SD)** | 84.7 (14.5) | 83.3 (14.0) | 67 524 | 0.053 |

*HRI-exercise = Heart Rate Increase during exercise phase, i.e. heart rate at peak exercise minus resting heart rate*
*HRD-recovery = Heart Rate Decrease during recovery phase. i.e. heart rate at peak exercise minus heart rate at recovery*
*PD = Parkinson's disease*

Results of the univariate cox regression analysis are presented in (Supplemental Table 4). The established prodromal clinical variables were significantly associated with incident PD. With regard to cardiac autonomic parameters, HRI-exc was associated with a 30% higher risk of incident PD per 10 beats less increase in heart rate during exercise. For HRD-rec, a blunted lowering in heart rate from peak exercise to recovery was associated with a 60% higher risk of incident PD per 10 beats difference.

In the minimally adjusted cox regression analysis, HRD-rec was significantly associated with incident PD, and HRI-exc was not (Table 3). After adjusting for clinical and autonomic prodromal factors, it was found that a blunted lowering in heart rate during recovery remained independently associated with a 30% higher risk of incident PD: HR 1.3; 95% CI 1.1-1.4; p < 0·001 per 10 beats less recovery (Table 3, Supplemental Table 5).

**Table 3.** Association of exercise heart-rate markers with incident PD, sequential adjustment for clinical and autonomic factors.

| Model | Marker | HR per 10 bpm (95% CI) | P Value |
| --- | --- | --- | --- |
| *Adjusted for age and sex* |  |  |  |
|  | HRI-ex | 1.0 (0.9 - 1.1) | 0.48 |
|  | HRD-rec | 1.3 (1.2 - 1.5) | <0.001 |
| *Adjusted for clinical variables <sup>a</sup>* |  |  |  |
|  | HRD-rec | 1.3 (1.1 - 1.5) | <0.001 |
| *Adjusted for clinical & prodromal autonomic variables <sup>b,c</sup>* |  |  |  |
|  | HRD-rec | 1.3 (1.1 - 1.4) | <0.001 |

*Hazard ratios are expressed per 10 beat/min impairment.*
*Abbreviations: HRI-ex = heart rate increase during exercise; HRD-rec = heart rate decrease during recovery.*
<sup>a</sup> *Clinical variables: age, sex, ethnicity, BMI, Townsend deprivation index, type-2 diabetes, and smoking.*
<sup>b</sup> *Prodromal autonomic variables: constipation, sleep duration, sleeplessness, depression, and bladder dysfunction.*
<sup>c</sup> *The interaction term between heart rate at peak-exercise and HRD-rec was not statistically significant ( $p = 0.53$ ).*

Figure 2 displays time-to-event curves. The median lag time to (hospital) diagnosis of incident PD was 9.3 years (IQR: 4.4.), showing the highest risk of incident PD in the subjects with the lowest HRD-red. After adjusting for clinical and autonomic prodromal factors, the association between HRD-rec and incident PD was found to be approximately log-linear (Figure 3).

**Figure 2.**
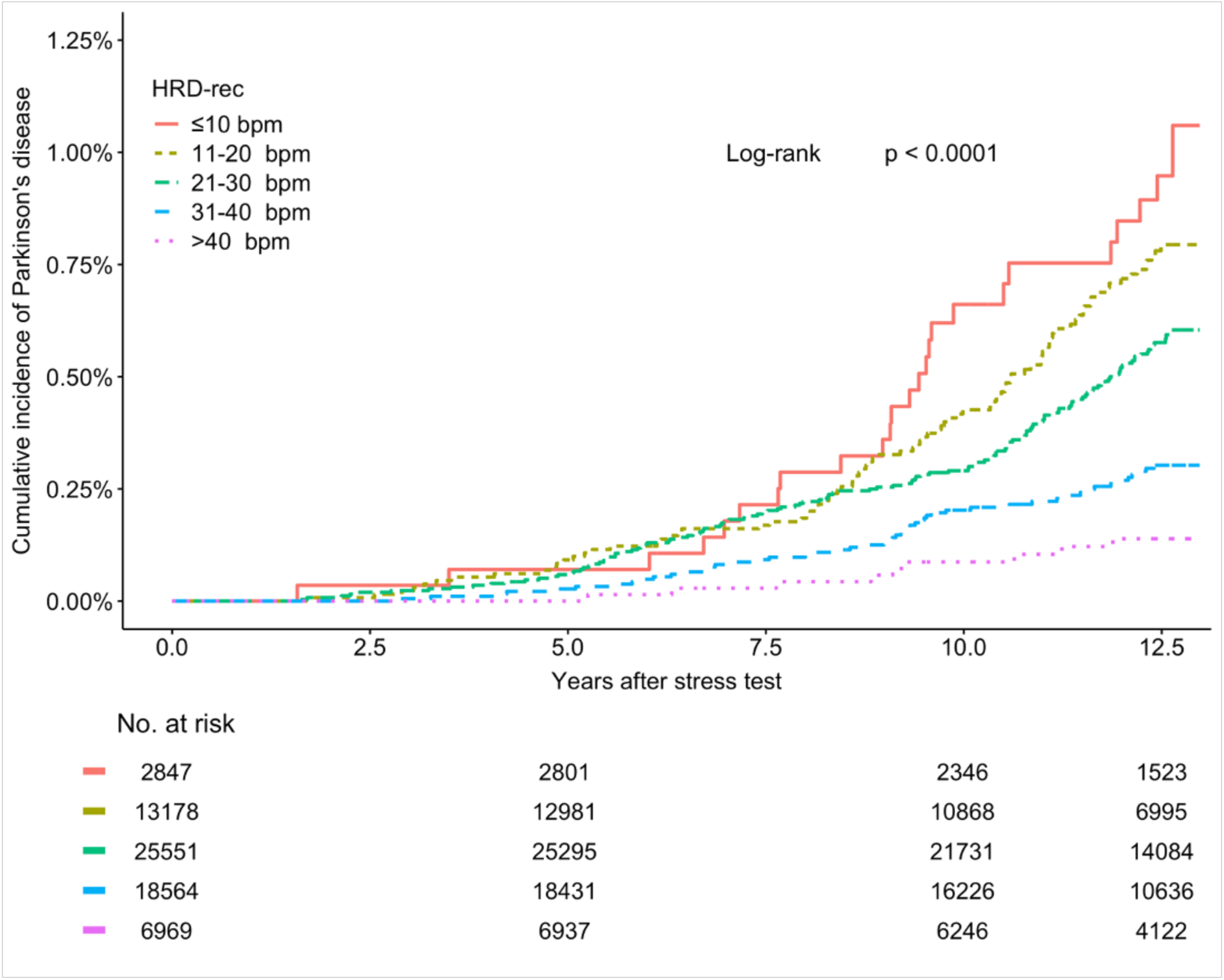
Unadjusted Kaplan-Meier estimates stratified by heart rate decrease during recovery (HRD-rec)

**Figure 3.**
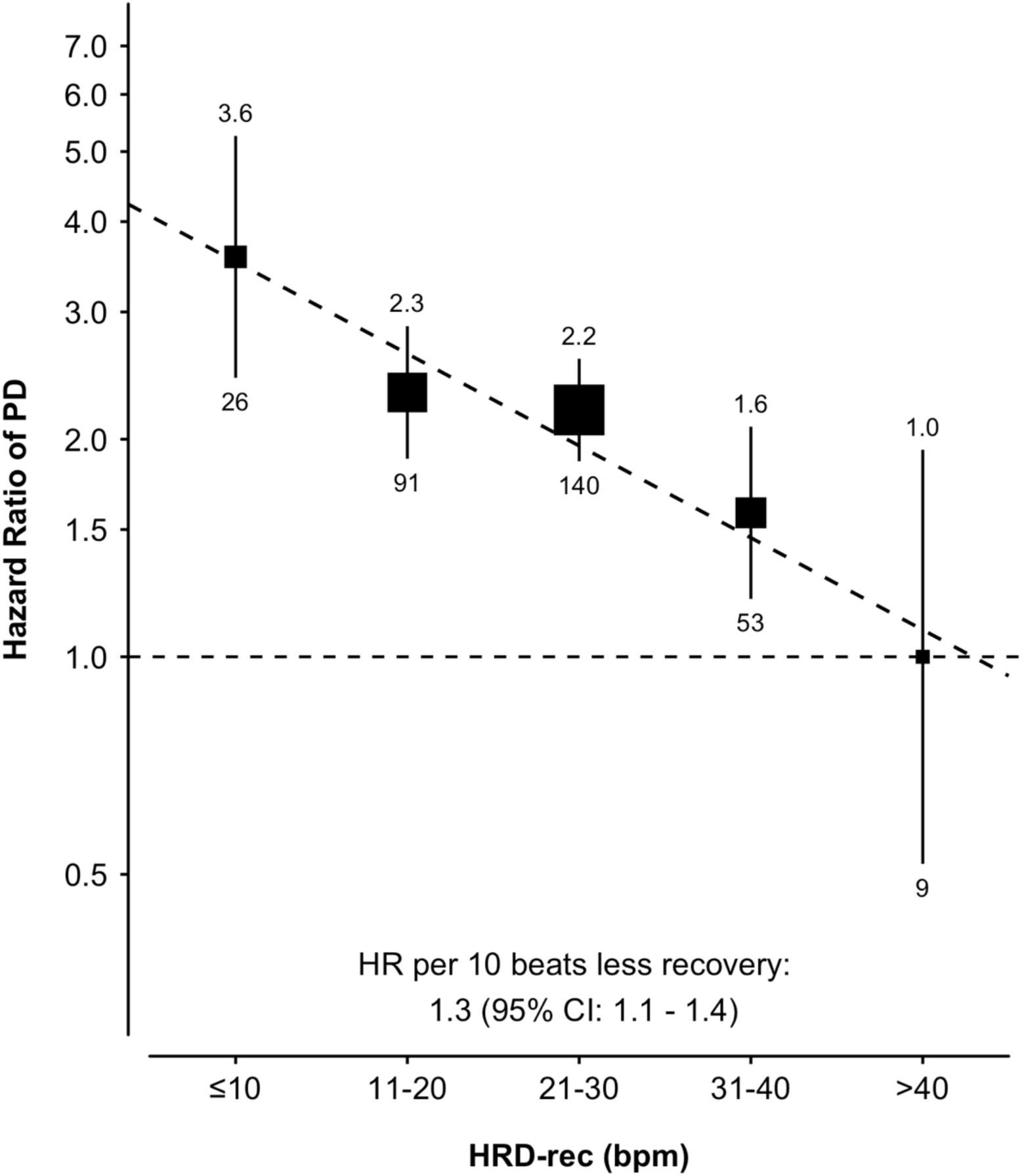
Association of heart rate decrease during recovery (HRD-rec) with risk of Parkinson’s disease. Adjusted for sex, age, ethnicity, Townsend deprivation index, body mass index, type-2 diabetes, smoking status, bladder dysfunction, constipation, depression, and self-reported sleep duration and sleeplessness; the number above each vertical line is the hazard ratio, and the number below is the number of incident cases.

### Sensitivity analyses

In both pre-specified sensitivity analyses with focus on medication, the association between HRD-rec and incident PD remained significant with similar risk estimates as the main analysis. In addition, findings and conclusions did not alter after the exclusion of individuals with imputed data (Supplemental Table 6).

Post-hoc sensitivity analyses focused on the association in relation to the achieved workload during the exercise test, and addressed the impact of imposing longer lag time to diagnosis. Outcomes did not alter the overall conclusions of the main analysis. Finally, we studied the association in the subset of participants with both out-of-hospital and in-hospital follow-up information, and findings were in line with the main analysis. (Supplemental Table 7)

## Discussion

The present report with 319 new cases of PD represents the largest population-based cohort to date on cardiac parameters of autonomic function as a potential prodromal marker, assessed by changes in heart rate during exercise and recovery. We found that an impaired capacity to reduce the heart rate during the recovery phase immediately after exercise was associated with incident PD, independently of established prodromal clinical and autonomic markers. Notably, our findings do not indicate a threshold value, but rather suggest a more graded association between a blunted heart rate recovery after exercise and incident PD (Figures 2 and 3). There was a univariate association between heart rate increase during the exercise phase and incident PD, which was no longer present after adjustment for sex and age. We also replicated earlier work by showing that subjects with prodromal parkinsonism had a greater likelihood of having established prodromal markers at baseline, including signs and symptoms indicative of non-cardiac autonomic dysfunction (i.e. constipation etc.). This suggests that the cohort studied here was representative of other prodromal populations described in the literature. Analogous to other established prodromal markers^23^, the lag time to diagnosis was long, with a median time of 9 years. This time interval represents the time until diagnosis made during an in-patient stay, so the actual lag time to a diagnosis established on an outpatient basis is shorter. Notably, while most prodromal markers are ‘static’ binary and non-modifiable variables (i.e. sex, hyposmia), we now present a candidate marker that is continuous and well known to be modifiable by interventions. Therefore, this marker has potential to be ‘dynamic’ as it may reflect quantitative changes over time by use of repeated measurements during follow-up. Longitudinal studies are warranted. As a quantifiable, continuous parameter, this cardiac autonomic marker may not only offer early diagnostic value, but also has promise to capture more subtle progressive changes during the prodromal phase. If proven to be modifiable in PD, it might be relevant when used as outcome measure in studies of putative disease-modifying interventions applied in the prodromal phase^24–26^.

In contrast to heart rate variability, that was studied in previous cohorts ^14,15^, heart rate recovery is a highly reproducible measure of parasympathetic activity^18,20^, with a straightforward protocol to register and calculate the parameter, using data contained within a standard exercise test. Heart rate recovery is also a recognized indicator of cardiovascular fitness^27^. The fact that participants who developed PD during follow-up versus those who did not show marked differences in other autonomic prodromal markers at the time of the exercise test (constipation, bladder dysfunction, etc) underscores that heart rate recovery in this subset of participants is a marker of altered autonomic function. This may well fit into the concept that the dorsal nucleus of the vagal nerve is one of the first manifestations in the neuropathological sequalae that lead towards clinically manifest motor PD^8,11,28^. Moreover, the association between HRD-rec and incident PD was similar in direction and magnitude when participants with a lower and higher workload were studied separately (Supplemental Table 6). With regard to previous cohort studies, the large Atherosclerosis Risk in Community (ARIC) cohort studied heart rate variability at rest and found an association with incident PD. During 20 years of follow-up in 12 162 patients (57% women, mean age 54 years), PD was diagnosed in 0·6% (78 cases)^14^. The smaller Cardiovascular Health Study (n= 5888) studied a community-based population of individuals who were 65 years and older (58% women, mean age 73 years). In this study heart rate variability was assessed during 24-hour Holter monitoring^15^. During 14 years of follow-up, PD was diagnosed in 3% of individuals, and in the subset (n=1587) with Holter monitoring no association was found between HRV and incident PD^15^. Although experimental studies have shown that HRV is sensitive to systemic cholinergic blockade^29^, the interpretation of HRV measures remains considerably more complicated than heart rate recovery, as it is strongly dependent on other physiological factors, including heart rate, and depth and frequency of respiration^29,30^. This aspect, together with non-uniformity between studies in the assessment of HRV (for example, 2 min resting ECG versus 24-hour registrations), hampers the interpretation of currently available evidence on HRV.

The increase in heart rate during exercise is primarily driven by sympathetic activation^18,19^. Both in patients with manifest motor PD, and in individuals in the prodromal phase, sympathetic involvement has been demonstrated^8,12,28,31^. A small case-control study on heart rate profiles during maximal exercise showed that the 18 studied cases had lower heart rates at peak exercise than the 36 matched controls, at a mean of 4 years prior to diagnosis. Heart rate recovery was not studied. A similar, but larger study did not detect significant signs of sympathetic dysfunction during the premotor phase of PD^17^.

In the present study, we observed that the heart rate increase during exercise was lower for individuals who developed PD during follow-up, but the association was no longer present after adjustment for sex and age, suggesting that sympathetic dysfunction is not an independent marker. Explanations for the discrepant findings with the small physiological study may lie in methodological aspects such as maximal versus submaximal exercise^32^, differences in sample size, or the selection of the incorporated confounding variables.

Our study is not without shortcomings. First, we acknowledge limitations in terms of generalizability with this rather healthy study cohort. Yet, with this particular study population, the association between heart rate recovery and incident PD is less likely to be flawed by co-morbidity. Second, the present work is a post-hoc analysis on a database where incident PD was not the primary outcome of interest. We therefore applied a strict case definition, that can be supported by source documents (a hospital admission confirming the ICD-code for PD). Notably, the hospital-based diagnosis was systematically collected during follow-up, in contrast to out-of-hospital diagnoses. The marked differences in baseline (prodromal) profiles between incident PD cases and the remainder of the group speaks for the solidity of our case definition, and the clinical representativeness of the cohort. Potentially, this case definition may have caused an underestimation of the actual incidence and may have influenced the effect estimations. Moreover, such a definition may result in a different case load in that patients with a hospital diagnosis may vary in clinical profile from those with an out-of-hospital diagnosis.

This is why we performed a post-hoc sensitivity analysis in the subset of participants with both information on the out-of-hospital and in-hospital phase using an alternative definition for incident PD, i.e. new out-of-hospital and/or in-hospital diagnosis of PD (Supplemental Table 7). The incidence of new cases was 0.5%, and results with regard to the association between HRD-rec and the new outcome measure were in line with the overall conclusions of the main analysis. With regard to the clinical profile of incident PD cases, we observed that in the aforementioned subset of participants, about 30% had an earlier out-of-hospital diagnosis. The diagnosis was made 2.4 years earlier than the in-hospital diagnosis. Appreciating that the majority of in-hospital diagnoses were first diagnoses, an interesting hypothesis is that these may have been more severe cases, as early involvement of body structures outside the nervous system (including the gut and the heart) is associated with a more malignant phenotype and a more rapid disease progression^12^.

We also acknowledge that the differential diagnosis from atypical parkinsonism (where the nature and extent of autonomic dysfunction is different) can be challenging, and a misclassification rate of up to 15% occurs in clinical studies of PD patients. However, such a misclassification would not have markedly affected the associations that we found, because early and prominent problems with cardiac innervation are not expected in most forms of atypical parkinsonism. Finally, our registrations did not meet the required duration and physical conditions for assessment of heart rate variability^29^, which unfortunately precludes comparisons with the previous cohorts^14,15^.

We foresee several areas for further research. First, our findings remain to be confirmed to provide the scientific background for initiatives to study its potential predictive value. To allow for a pooled comparison across studies that tested the same autonomic parameter, it is imperative to focus on better standardization of assessment protocols^29,30^. Second, with the growing notion of distinct patterns in the development towards PD, defining a brain-first and a body-first subtype^12^, more detailed studies are warranted in these subpopulations^24,25^. It is plausible that the potential diagnostic value will be greater in the body-first subtype, where the autonomic nervous system is involved much earlier in the disease process.

Taken together, our findings provide additional supportive evidence for the concept that pathophysiological changes in the autonomic nervous system may precede the clinically manifest motor syndrome of PD. If confirmed in an independent cohort, we anticipate that autonomic cardiac biomarkers could be incorporated in a more comprehensive battery of early diagnostic biomarkers^5,33^. Potentially, as a quantitative, potentially modifiable marker, it may pave the way for longitudinal biomarker studies on therapeutic interventions.

## Acknowledgements

The Center of Expertise for Parkinson’s Disease & Movement Disorders was supported by a centre of excellence grant of the Parkinson’s Foundation. SvD and AD are funded by the Wellcome Trust [223100/Z/21/Z] and the British Heart Foundation Centre of Research Excellence. For the purpose of open access, the author(s) has applied a Creative Commons Attribution (CC BY) license to any Author Accepted Manuscript version arising.

## Author Contributions

S.vD, M.B., B.B., A.D., contributed to the conception and design of the study, S.vD, M.B., J.R., M.O., J.I., B.B., A.D. contributed to the acquisition and analysis of data; S.vD, M.B., J.R. J.S. S.D, M.O., A.T., P.M. J.T., L.E., J.I., P.L., B.B, and A.D. contributed to drafting the text and preparing the figures.

## Data availability statement

Data from the UK Biobank are available to researchers on application to the UK Biobank (https://www.ukbiobank.ac.uk/enable-your-research/about-our-data).

