## Supplemental Tables for "Cardiac autonomic function during exercise and incident Parkinson’s disease"

**Supplement Table 1: Definitions of covariates and diseases variables in UK Biobank**

| | HES statistics ICD-10 and ICD9 codes | HES statistics OPCS4 codes | Self-reported Data-Field 20002 | Self-reported operations Data-Field 20004 | Other data fields |
|---|---|---|---|---|---|
| Type-2 diabetes | E10, E11, E13, E14 | | 1223 | | Field 2443 = 1 |
| Constipation | K590 | | 1599 | | Field 6154 = 6 |
| Depression | F33 | | 1286 | | |
| Bladder dysfunction | N31 | | 1202 | | |
| Sleep duration | | | | | Field 1160 |
| Sleeplessness | | | | | Field 1200 = 3 |
| Erectile dysfunction | | | 1518 | | |
| Parkinson's disease | G20 | | 1262 | | |
| ***Prevalent neurodegenerative disease*** | | | | | |
| Extrapyramidal and movement disorders | G20-G26 | | 1262 | | |
| Other degenerative diseases of the nervous system | G30-G32 | | | | |
| Multi-system degeneration of the autonomic nervous system | G903 | | | | |
| ***Prevalent Cardiovascular disease*** | | | | | |
| Ischemic Heart Disease | 411.1, 410.11, 410.41, 410.81, 410.91, 410.71, 410.01, 410.21, 410.31, 410.51, 410.61, 411.89, 429.2, 414.0, 414.10, 414.19, 414.8, 414.9, I20-I25 | K40, K41, K42, K44, K45, K49, K50, K75 | | 1070, 1095, 1523 | |
| Cardiomyopathies | 425.4, 425.11, 425.18, 425, 425.3, 425.5, 425.9, 425.2, 425.8, 425.7, I42-I43 | | | | |
| Heart Failure | 428.0, 428.1, 428.9, I13.0, I13.2, I50 | K59.6, K61.7, K60.7 | | | |
| Arrhythmias | 427.3, 427.31, 427.32, 427.1, 427.9, 427.5, I48, I47.2, I47.0, I49.0, I46.0, I46.1, I49.0, I44.1, I44.2, I49.9, G45 | K576, K641, X504, X504 | | | |
| Stroke | 434.91, 434.01, 434.11, 436, I63, I61, I64 | | | | |
| Other cardiovascular diseases | I70, I73, I74, I71, I11, I34, I35, I36, I37, Q20, Q21, Q22, Q23, Q24, B33.2, I40, I41, I51.4 | K59, K72 | | | |
| ***Active cancer diagnosis*** | | | | | |
| Malignant neoplasms | C00-C97 | | | | |

**Supplemental Table 2**
**Characteristics of UK Biobank participants with and without consent for the exercise test**

| | **Consent for exercise test** | | | |
|---|---|---|---|---|
| | **Yes (n=96 524; 19.2%)** | **No (n=405 840; 80.8%)** | **N** | **p** |
| Sex, male, % | 46.1% | 45.5% | 502 364 | <0.001 |
| Age, years, mean (SD) | 56.9 (8.0) | 56.4 (8.1) | 502 364 | <0.001 |
| Ethnicity, white, % | 92.1% | 95.2% | 499 587 | <0.001 |
| BMI, mean (SD) | 27.3 (4.8) | 27.5 (4.8) | 499 259 | <0.001 |
| Townsend deprivation index[a], mean (SD) | -1.2 (3.0) | -1.3 (3.1) | 501 740 | <0.001 |
| Type 2 Diabetes Mellitus, yes, % | 5.5% | 5.4% | 502 364 | 0.13 |
| Current Smoking, yes, % | 9.1% | 11.0% | 499 415 | <0.001 |
| Constipation, yes, % | 3.7% | 3.8% | 502 364 | 0.38 |
| Depression, yes, % | 5.9% | 6.1% | 502 364 | 0.017 |
| Bladder dysfunction, % | 0.9 % | 0.5 % | 502 364 | <0.001 |
| Erectile dysfunction, % | 0.4 % | 0.1 % | 229 067 | <0.001 |
| Sleep duration, hours, mean (SD) | 7.1 (1.2) | 7.1 (1.3) | 501 476 | 0.025 |
| Sleeplessness, % | 27.0% | 28.6% | 499 984 | <0.001 |

There are statistically significant but clinically irrelevant differences in baseline characteristics between the 96,524 individuals who provided consent for the exercise test, and the remaining part of the UK Biobank cohort. *BMI = Body Mass Index, SD = standard deviation.*

[a] *Townsend deprivation index is a is a measure of material deprivation.*

**Supplemental Table 3**
**Characteristics of participants consenting for the exercise test in relation to eligibility for present study**

| | Included in study | | | |
|---|---|---|---|---|
| | **Yes (n=69 288; 71.8%)** | **No (n=27 236; 28.2%)** | **N** | **p** |
| Sex, male, % | 47.6% | 42.4% | 96 524 | <0.001 |
| Age, years, mean (SD) | 56.0 (8.0) | 59.1 (7.5) | 96 524 | <0.001 |
| Ethnicity, white, % | 93.1% | 89.5% | 95 896 | <0.001 |
| BMI, mean (SD) | 27.0 (4.4) | 28.2 (5.5) | 96 178 | <0.001 |
| Townsend deprivation index[a], mean (SD) | -1.4 (2.9) | -0.8 (3.2) | 96 420 | <0.001 |
| Type 2 Diabetes Mellitus, yes, % | 4.0% | 9.5% | 96 524 | <0.001 |
| Current Smoking, yes, % | 8.5% | 10.5% | 95 938 | <0.001 |
| Constipation, yes, % | 2.9% | 5.8% | 96 524 | <0.001 |
| Depression, yes, % | 5.2% | 7.8% | 96 524 | <0.001 |
| Bladder dysfunction, % | 0.8% | 1.4% | 96 524 | <0.001 |
| Erectile dysfunction, % | 0.4% | 0.5% | 44 498 | 0.032 |
| Sleep duration, hours, mean (SD) | 7.1 (1.1) | 7.1 (1.5) | 96 308 | 0.52 |
| Sleeplessness, % | 24.8% | 32.8% | 96 053 | <0.001 |

*BMI = Body Mass Index, SD = standard deviation. [a] Townsend deprivation index is a is a measure of material deprivation.*

**Supplemental Table 4**
**Hazard ratios of characteristics at baseline – univariate associations with incident PD**

| | **Hazard Ratio (95% CI)** | **p** |
|---|---|---|
| Sex, male | 2.5 (2.0-3.2) | <0.001 |
| Age, per year increase | 1.1 (1.1-1.2) | <0.001 |
| Ethnicity, white | 1.6 (1.0-2.7) | 0.070 |
| Townsend deprivation index[a], per unit increase | 1.0 (0.9-1.0) | 0.039 |
| BMI, per unit increase | 1.0 (1.0-1.1) | 0.016 |
| Type 2 Diabetes Mellitus, yes | 3.6 (2.6-4.9) | <0.001 |
| Current Smoking, yes | 0.4 (0.2-0.7) | 0.001 |
| Constipation, yes | 2.1 (1.3-3.3) | 0.002 |
| Depression, yes | 1.7 (1.2-2.5) | 0.006 |
| Bladder dysfunction, yes | 1.8 (0.8-4.1) | 0.15 |
| Sleep duration, per hour increase | 1.2 (1.1-1.3) | 0.004 |
| Sleeplessness, yes | 0.9 (0.7-1.2) | 0.40 |
| HRI-exc, per 10 beats less increase | 1.3 (1.2-1.4) | <0.001 |
| HRD-rec, per 10 beats less recovery | 1.6 (1.4-1.7) | <0.001 |

*[a] Townsend deprivation index is a is a measure of material deprivation.*
*BMI = Body Mass Index*
*HRI-exc = Heart rate increase during exercise*
*HRD-rec = Heart rate decrease during recovery*

**Supplemental Table 5**
**Associations of clinical and autonomic exercise parameters with incident PD: multivariate analysis**

| | Hazard Ratio (95% CI) | p |
|---|---|---|
| Sex, male | 2.3 (1.8-2.9) | <0.001 |
| Age, per year increase | 1.1 (1.1-1.2) | <0.001 |
| Ethnicity, white | 1.0 (0.6-1.8) | 0.88 |
| BMI, per unit increase | 1.0 (1.0-1.0) | 0.86 |
| Townsend deprivation index[a], per unit increase | 1.0 (1.0-1.0) | 0.98 |
| Type 2 Diabetes Mellitus, yes | 2.2 (1.6-3.1) | <0.001 |
| Current Smoking, yes | 0.4 (0.2-0.8) | 0.005 |
| Constipation, yes | 2.1 (1.3-3.3) | 0.003 |
| Depression, yes | 2.0 (1.4-3.0) | 0.001 |
| Bladder dysfunction, yes | 1.2 (0.5-2.7) | 0.65 |
| Sleep duration, per hour | 1.0 (0.9-1.2) | 0.38 |
| Sleeplessness, yes | 0.8 (0.6-1.1) | 0.17 |
| HRD-rec, per 10 beats less recovery | 1.3 (1.1-1.4) | <0.001 |

*[a] Townsend deprivation index is a is a measure of material deprivation.*
*BMI = Body Mass Index,*
*HRD-rec = Heart rate decrease during recovery phase after exercise*

**Supplemental Table 6**
**Adjusted hazard ratios of incident PD according to heart rate decrease during recovery in prespecified sensitivity analyses**

| Pre-specified sensitivity analyses | | | |
|---|---|---|---|
| Marker | N | HR per 10 bpm (95% CI) | P Value |
| **Original analysis** | | | |
| HRD-rec | 69,288 | 1.3 (1.1 - 1.4) | <0.001 |
| **Exclusion of individuals using heart rate modulating medication** | | | |
| HRD-rec | 65,802 | 1.3 (1.1 – 1.5) | <0.001 |
| **Exclusion of individuals using dopaminergic and/or (other) psychoactive medication** | | | |
| HRD-rec | 64,578 | 1.3 (1.2 - 1.5) | <0.001 |
| **Exclusion of individuals with missing data** | | | |
| HRD-rec | 66,282 | 1.3 (1.1 - 1.4) | <0.001 |

*Medication data (UK Biobank Data Field: 20003) were mapped to the Anatomical Therapeutic Chemical (ATC) classification system (https://github.com/PhilAppleby/ukbb-srmed).Heart rate modulating medication: beta and calcium blockers, ATC codes: C07, C08; psychoactive medication: ATC codes: N04, N05, N06, N07. Analyses were adjusted for both clinical variables (age, sex, ethnicity, body mass index (BMI), Townsend deprivation index, type-2 diabetes, and smoking) and prodromal autonomic variables (constipation, sleep duration, sleeplessness, depression, and bladder dysfunction).*

*HRD-rec = heart rate decrease during recovery. Hazard ratios (HR) and confidence intervals (CI) are expressed per 10 beat/min impairment*

**Supplemental Table 7**
**Adjusted hazard ratios of incident PD according to heart rate decrease during recovery in post-hoc sensitivity analyses**

| Post-hoc sensitivity analyses | | | |
|---|---|---|---|
| **Analysis stratified by workload[a]** | | | |
| **HRD-rec<br>only considering subjects exercising at 50% of maximum predicted workload** | 58,728 | 1.3 (1.1 – 1.5) | 0.001 |
| **HRD-rec<br>only considering subjects exercising at 35% of maximum predicted workload** | 8381 | 1.2 (0.8 – 1.6) | 0.829 |
| **Imposing longer lag time to diagnosis** | | | |
| **HRD-rec<br>only considering cases after median follow-up time (9.3 yrs)** | 67,252 | 1.3 (1.1 – 1.5) | 0.007 |
| **HRD-rec<br>only considering cases after $75^{th}$ % of follow-up time (11.0 yrs)** | 54,417 | 1.2 (0.9 – 1.5) | 0.163 |
| **Incident PD defined as new out-of-hospital and/or in-hospital diagnosis of PD[b]** | | | |
| **HRD-rec** | 37,458 | 1.2 (1.0 – 1.5) | 0.013 |

[a] *The interaction term between HRD-rec and workload (35% or 50% of maximum predicted workload) was p = 0.548.*

[a] *Incident PD defined by self-reported data (see Supplemental Table 1), primary care data (diagnostic Read code F12..00 for Parkinson's disease), and/or hospitalization (ICD 10 G20); there were 171 cases (0.5%). Median time to out-of-hospital diagnosis was 4.07 years (IQR: 2.25 – 5.09).*
